# COVID-19: Mechanistic model calibration subject to active and varying non-pharmaceutical interventions

**DOI:** 10.1101/2020.09.10.20191817

**Authors:** Mark J. Willis, Allen Wright, Victoria Bramfitt, Victor Hugo Grisales Díaz

**Affiliations:** School of Engineering, Newcastle University, Newcastle upon Tyne, NE1 7RU, UK; Grupo de investigación en Microbiología y Biotecnología MICROBIOTEC, Facultad Ciencias de la Salud, Universidad Libre Seccional Pereira,. Belmonte Avenida Las Américas

**Keywords:** COVID-19, model calibration, discrete event modelling and management, management, time varying disease transmission rate

## Abstract

Mathematical models are useful in epidemiology to understand the COVID-19 contagion dynamics. Our aim is to demonstrate the effectiveness of parameter regression methods to calibrate an established epidemiological model describing COVID-19 infection rates subject to active and varying non-pharmaceutical interventions (NPIs). To do this, we assess the potential of some established chemical engineering modelling principles and practice for application to modelling of epidemiological systems. This allows us to exploit the sophisticated functionality of a commercial chemical engineering simulator capable of parameter regression with piecewise continuous integration and event and discontinuity management. Our results provide insights into the outcomes of on-going disease suppression measures, while visualisation of reported data also provides up-to-date condition monitoring of the status of the pandemic. We observe that the effective reproduction number response to NPIs is non-linear with variable response rate, magnitude and direction.

**Highlights:**

- Modelling COVID-19 contagion dynamics with piecewise continuous integration and event and discontinuity management
- Equivalence to kinetic model with time varying stoichiometry
- Model calibration and estimation of non-linear variation in *R_e_*
- Development of prototype demonstrator algorithm for estimation of *R_e_* using data with time varying NPIs

## 1.0 Introduction

COVID-19 is currently a global pandemic affecting around 213 countries around the world. As of 31 August 2020, 25.6million cases, with 17.9 million recovered patients and 859,550 deaths have been reported [1]. To control the pandemic, most governments have issued recommendations such as intensified hand hygiene and have taken measures such as closing borders, enforcing lockdowns, etc. These NPIs reduce infection rates, keeping the number of severe cases below hospital capacity limit, a strategy popularly referred to as ‘flattening the curve’. A significant challenge is to identify and efficiently evaluate the effect that active and varying NPIs have on the disease transmission rate. This is particularly important as countries begin to relax NPIs after successfully flattening the curve of active cases.

### 1.1 The effective reproduction number

Key parameters used to quantify contagion are the basic and effective reproduction numbers. These dimensionless numbers describe the average number of expected secondary infections generated by each infected person in the absence and presence of controlled interventions. Current opinion suggests that the COVID-19 has a basic reproduction number ∼2 — 3. Although a recent review [2] compared twelve studies published from the 1st of January to the 7th of February 2020 which reported a range of values for the COVID- 19 basic reproduction number between 1.5 and 6.68. This apparent disparity arises because the reported number depends on country, culture, the stage of the outbreak and calculation method used. NPIs aim to slow the spread of the virus and reduce the effective reproduction number to a sustained value less than one so that the pandemic will eventually die out. Scientists and governments in many countries around the world use the effective reproduction number as an illustrative metric to explain and justify the introduction and relaxation of NPIs [3].

### 1.2 Mathematical modelling

Most reported effective reproduction numbers are estimates obtained from mathematical models. These include mechanistic transmission models [4], statistical models, [5], [6] deterministic epidemiological models [7] – [9] and a statistical dynamical growth model [10]. Estimated values of the effective reproduction number are highly dependent on the choice of the model, the initial conditions used to parameterise the model as well as underlying model assumptions.

A widely used compartmental model in epidemiology is the susceptible – infected – removed (*SIR*) model, [11] – [13]. An extension of this model separates the removed group into recovered and dead (*SIRD*). These models differentiate between the classes of individuals, modelling transition rates between the classes using rate laws defined in accordance with the law of mass action kinetics. As noted in [14], modelling rates of infection through analogy to chemical kinetics is the standard approach in mathematical epidemiology. Indeed, the purpose of mathematical modelling of epidemiology is mostly concerned with the kinetics of the spread of a contagion, which is clearly an important issue when epidemics occur. The (*SIR*) and (*SIRD*) models are deterministic, autocatalytic kinetic models of the whole population [15].

### 1.3 Capturing essential system dynamics

Any mathematical model must capture essential system dynamics for calibration, predictive modelling and simulation studies to be meaningful. For COVID-19 and the application of NPIs, these are an initial exponential growth in active cases, slowing as the NPIs influence disease transmission. After a peak in the number of new cases, there will typically be a slow decline in active cases, provided the NPIs are not excessively relaxed.

While the *SIR* model and its extensions provide the fundamental backbone to represent these dynamics, there is no accepted means to alter the disease transmission rate in order to ‘flatten-the curve’. For example, recent studies used a deterministic model with a constant reproduction number to model the outbreak dynamics in Europe [16] and in China [17] by using a reduction in the total population as a model calibration parameter that indirectly quantifies the application of NPIs. Whereas [18] use a hyperbolic tangent function to capture time variation in the effective reproduction number and [19] adjust model rate constants assuming a sigmoidal profile with respect to time. Our recent work [20] demonstrates that the *SIR* model augmented with a differential equation to model decline in the disease transmission rate could accurately model reported case data, and hence determine the effective reproduction number. However, in [20] the effective reduction in disease transmission rate was characterised because of the application of all NPIs. Moreover, model calibration and validation did not utilise the most recently available case data as NPIs have begun to be relaxed. In this paper, we extend previous work to model variations in the effective reproduction number using a philosophy centred on discrete event modelling and management in order to calibrate the *SIRD* model to reported case data with active and varying NPIs.

### 1.4 Exploiting the functionality of commercial dynamic modelling with kinetic fitting tools

The *SIRD* epidemiological model is structurally equivalent to the model equations for a set of chemical reactions occurring in a well-mixed batch reactor in which the stoichiometry of the contagion reaction varies. Due to the structural equivalence of the chemical and epidemiological models, we can exploit the sophisticated functionality of a commercial chemical engineering simulator that combines a dynamic modelling framework with kinetic regression tools.1 The software platform provides an environment for rapid development of piecewise continuous models containing a series of discrete operational events. We adapt existing functionality for sequencing of discrete events to represent NPIs and characterise the efficacy of NPIs on reducing and maintaining the effective reproduction number to acceptable levels. This allows us to calibrate the characteristic rates of infection and removal of individuals and estimate the effective reproduction number throughout the epidemic. Further, a successfully calibrated model allows dynamic simulation studies to quantify the effect of the relaxation of the NPIs.

## 2.0 Methods

Up-to-date daily information regarding the number of active, recovered and fatal cases for most countries around the world is available from [1]. Retrieving this information allows daily model calibration, on a country-by-country basis, incorporating information about the timing and nature of their NPIs as well as the monitoring of the status of the pandemic.

### 2.1 Kinetic modelling applied to an epidemiological system

Defining *I* as an infected individual, *R* as a recovered individual and *D* as a deceased individual, the stoichiometric scheme describing the transition of individuals between the four compartments of an *SIRD* model is

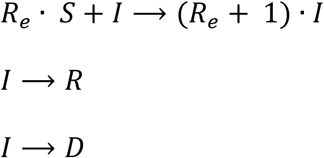

In this scheme, *R_e_* is the (dimensionless) effective reproduction number. We observe that it is analogous to a stoichiometric coefficient in a chemical reaction scheme. The significant difference is that in chemical schemes, stoichiometric coefficients are constant whereas the effective reproduction number, *R_e_* can vary throughout the course of an epidemic as NPIs are applied.

We can develop a set of model equations for this epidemiological scheme by treating it in the same way as a chemical scheme and applying the law of mass action^2^. Defining, *n_S_* (people) as the number of susceptible *n_I_* (people) the number of infected, *n_R_* (people) the number of recovered, *n_D_* (people) the number of deceased, the rate of change of the number of people in the various compartments of the model are,

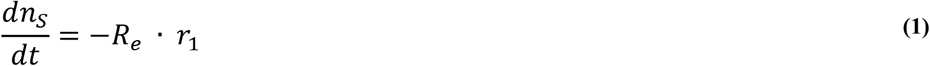

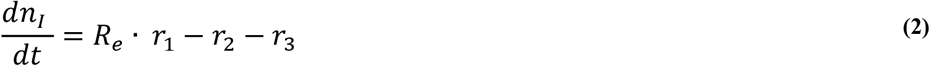

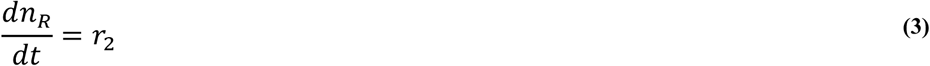

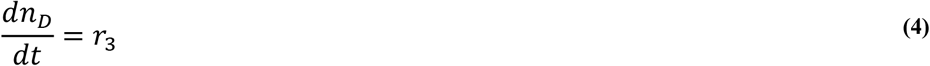

The rate terms *r*_*i*,(*i* = 1,2,3)_(people ▪ day^−1^) are,

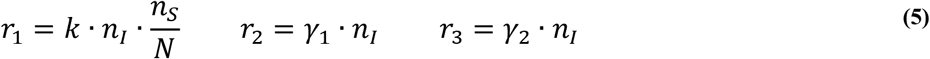

In equations (5), *k* (day^−1^) is the infection rate constant, *γ*_1_ (day^−1^) is the removal rate constant of recovered infectious individuals, *γ*_2_ (day^−1^) is the removal rate constant of deceased individuals, *N* (people) is the total population. Substituting these rate terms into(1) to (4) gives,

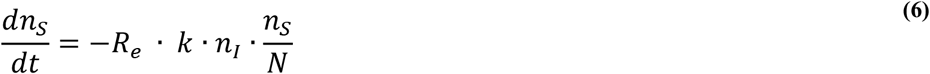

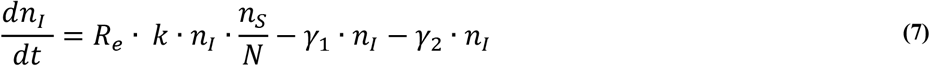

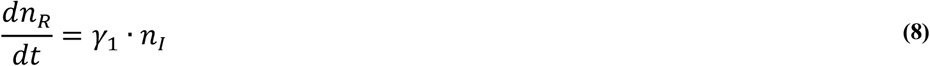

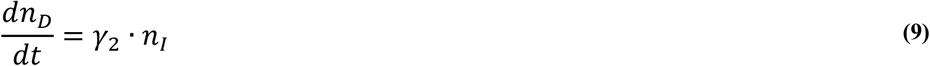

### 2.2 Modelling the variation in the effective reproduction number

In equations (6) – (9) we assume that *k* is constant and refer to this as the specific transmission probability per exposure time, a constant that is characteristic of the COVID-19 infection. It is assumed that the effective reproduction number *R_e_* varies as an exponential function, and that the variation of *R_e_* is due to measures taken as NPIs are changed during the epidemic. We use an Arrhenius equation to represent the exponential variation,

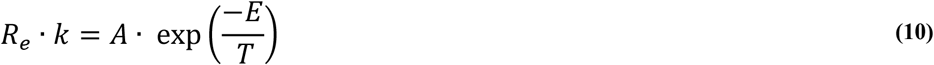

In equation (10), *A* and *E* are constants determined by model calibration at the same time as the other unknown parameters in equations (6) – (9). In order to use this equation to capture variation in *R_e_* we use temperature as a placeholder variable to represent the efficacy of NPIs. This allows us to exploit the discrete event based operational modelling capability in a commercial chemical engineering simulator and to introduce a sequence of temperature changes to emulate the effect of NPIs.

### 2.3 Using a commercial simulation package to construct the model

In this study, we exploit the features of an existing simulator designed for regression of chemical kinetic rate constants and predictive modelling of chemical systems. We use this simulator as a demonstrator to model an epidemiological system and calculate of variation in the effective reproduction number in response to NPIs. We enter the *SIRD* reaction scheme including the reaction rate terms (5) directly into the software. The software automatically constructs the associated set of ODEs (6) – (9). Next, we specify operational information such as the initial *SIRD* numbers. All models are constructed as batch operation, although we note that the simulator’s capability for fed batch operation would allow modelling of influx of infected cases to the population.

The model also includes a sequence of temperature steps and ramps, which we use to represent the NPIs. This sequence is shown conceptually in Figure 1.

**Figure 1.**
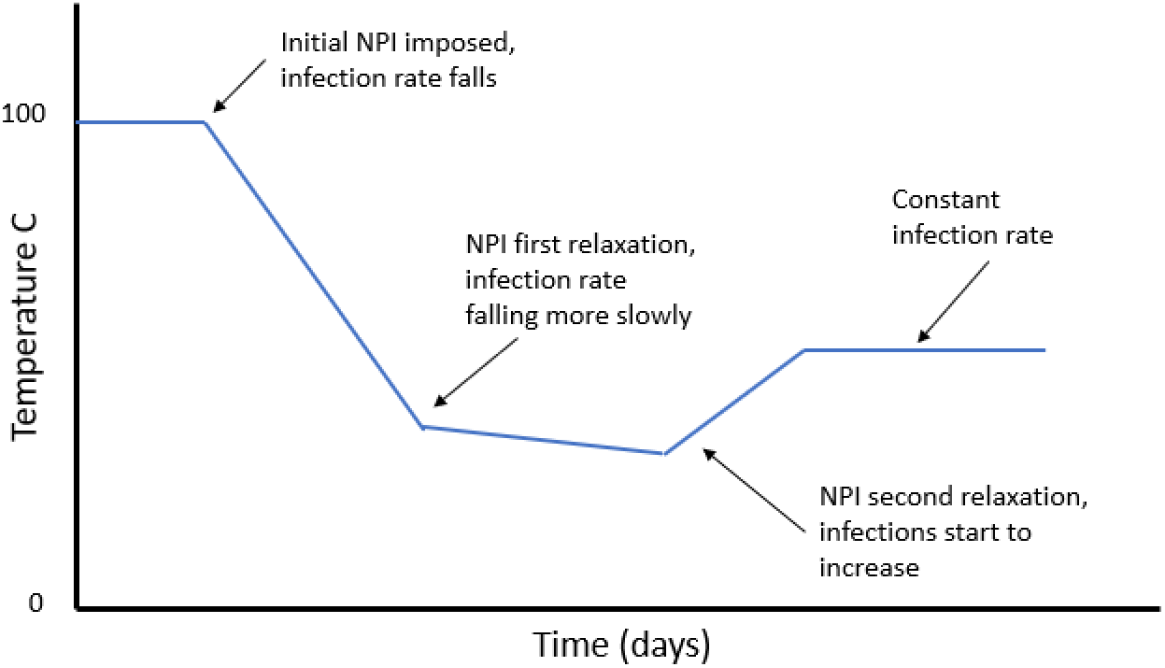
Schematic of a typical temperature profile used.

Nominally, we use an initial baseline temperature value of 100°C to represent the system before any interventions are applied. To represent the introduction of NPIs, we impose a negative temperature ramp that can vary in rate and duration. As NPIs are changed the temperature can be further manipulated to reflect the observed change in active cases. The magnitude, direction and rate of the subsequent temperature changes in the model is representative of the stringency and efficacy of the NPIs. Increasing stringency of NPIs represented by decreasing temperature, relaxation of NPIs by increasing temperature. Interpreting temperature as the efficacy of applied NPIs provides a quantitative measure of their effect and insight into the dynamics of the disease.

### 2.4 Model calibration

The modelling software used in this study is designed for simulation studies of batch chemical systems. To calibrate our model, we must calculate the kinetic constants and the values for the temperature profile used as a placeholder for the NPIs.

The simulator has methods for regression of kinetic constants, but does not have a capability for optimisation of a temperature profile. Therefore, to perform model calibration we combine the existing kinetic regression capability in the simulator with systematic manual manipulation of the temperature sequence representing NPIs. These manual interventions can be considered as a prototype demonstrator of the algorithmic steps required to develop tools for modelling epidemiological systems.

The process kinetic constants are manipulated by an optimisation algorithm in the simulator to vary the pre-exponential term and exponent for R_e_ ▪ *k* as well as the rate constants *γ*_1_ and *γ*_2_. This is performed with a data window over the duration of the first NPI. The kinetic constants calculated in this initial phase are held constant for the remaining model calibration phases. In subsequent model calibration phases, when observation of the predicted number of infected persons shows a divergence from the recorded data after a period-of-time (corresponding to a change in the initial NPI) the data window is extended and the next section of the temperature profile is determined by manual intervention, such that the objective function for the data window is minimised and a good fit to data for the whole of the data set in use is maintained.

To quantify the discrepancy between model response and reported case data, the sum of the squared error between the reported cumulative infected individuals *n_I_(t)* and the model prediction *n_I_^∗^(t)* as well as the reported removed individuals, *n_R_(t)* and the model prediction *n_R_^∗^(t)* and the reported dead *n_D_(t)* and the model prediction *n_D_^∗^(t)* is calculated as,

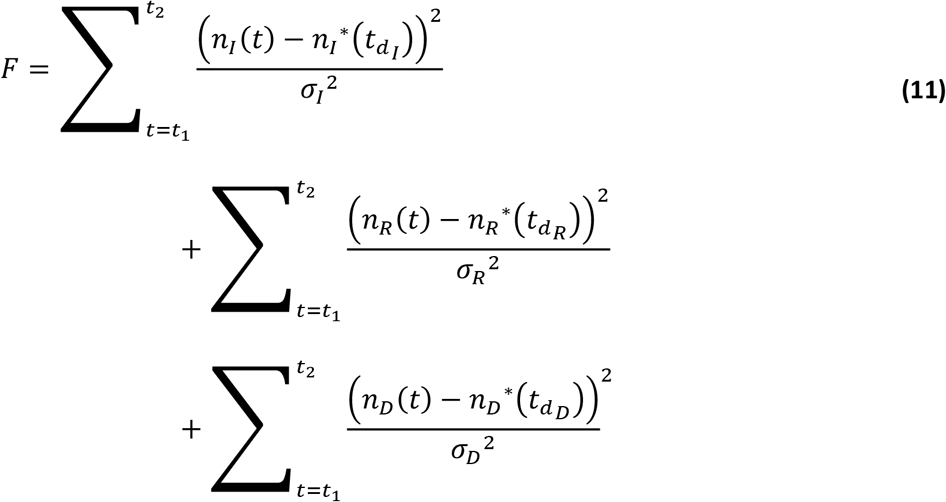

The sum is over the time-period *t*_1_ (the initial outbreak reaching exponential growth) and *t*_2_ (current time). *σ*^2^ is the variance in the reported data for a given data set. The parameter *t_d_* represents a time delay associated with the reporting of recovered / deceased individuals. This delay accounts for the time it takes for confirmation of deaths, recoveries, or the validation of data from tests for infection.

The optimisation algorithm used for model calibration was a modified Simplex algorithm^3^ and numerical integration of the ODEs is via an adaptive Runge-Kutta integrator. The integration algorithm is piecewise continuous with event and discontinuity management. This ensures accurate model response to interventions, for example the sequence of NPIs as represented by the temperature profile.

The temperature profile is specified manually for each country. For all the models developed, the initial temperature is set to an arbitrary initial value of 100°C. We use known intervention times from reports of actions taken by governments. The initial decrease in temperature commences with the introduction of the first NPIs. The time and temperature values for the end-point of an initial linear temperature ramp are adjusted manually such that the trajectories of the model predictions correspond with the reported data for the early stage of the epidemic extending beyond the peak in the number of infected cases. The rates of change and durations of the subsequent changes in the temperature profile are adjusted empirically as the model is calibrated such that a good fit to the entire data set is achieved.

### 2.5 Calculating the effective reproduction number

The number of infected individuals passes through a maximum at *t_I_max__*, and at this point

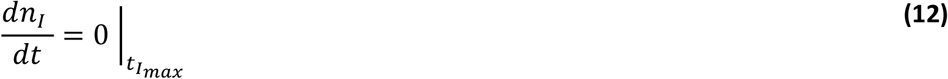

It follows that

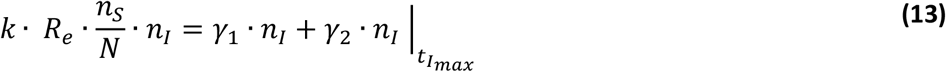

If the effective reproduction number at this point is, *R_e_* = 1, then the constant for specific transmission probability per exposure time can be calculated as

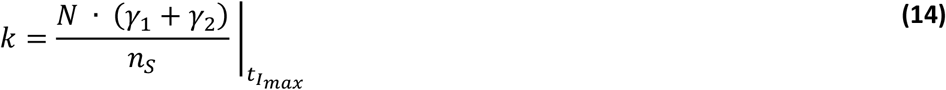

The value of the placeholder variable *T*(*t*) is known by inspection of the temperature profile imposed on the model. This allows the instantaneous effective reproduction number *R_e_*(t) to be calculated using (10).

## 3.0 Results

To demonstrate and discuss our modelling, we use reported case data from Germany, Austria, Saudi Arabia and Italy. As with any data-driven approach, it is only possible to have representative and reliable results if the data is also reliable. The countries selected have a reasonably well developed COVID-19 testing capacity (test coverage is greater than 15 per thousand residents), which would imply reasonably robust and reliable data. The time period we consider is from around the 1^st^ March to 31^st^ August, 2020.

Given our model formulation, a change in temperature translates directly into changes in the effective reproduction number. Imposing a linear decrease in temperature results in an exponential decrease in *R_e_* with time. A constant temperature input to the model results in a constant output of *R_e_*. The dynamic response to an NPI is represented by the rapidity of the temperature change, and the variation in *R_e_* correlates with the variation in the temperature placeholder variable.

### 3.1 Germany

Figure 2 shows *SIRD* model predictions (***R*_squared_** = 99.9%) plotted with the reported values for numbers of active infected, recovered and deceased individuals. The time shifts for reporting delays are *t_dI_* = 0, *t_d_R__* = 6 *days, t_d_D__* = 6 *days*.

**Figure 2.**
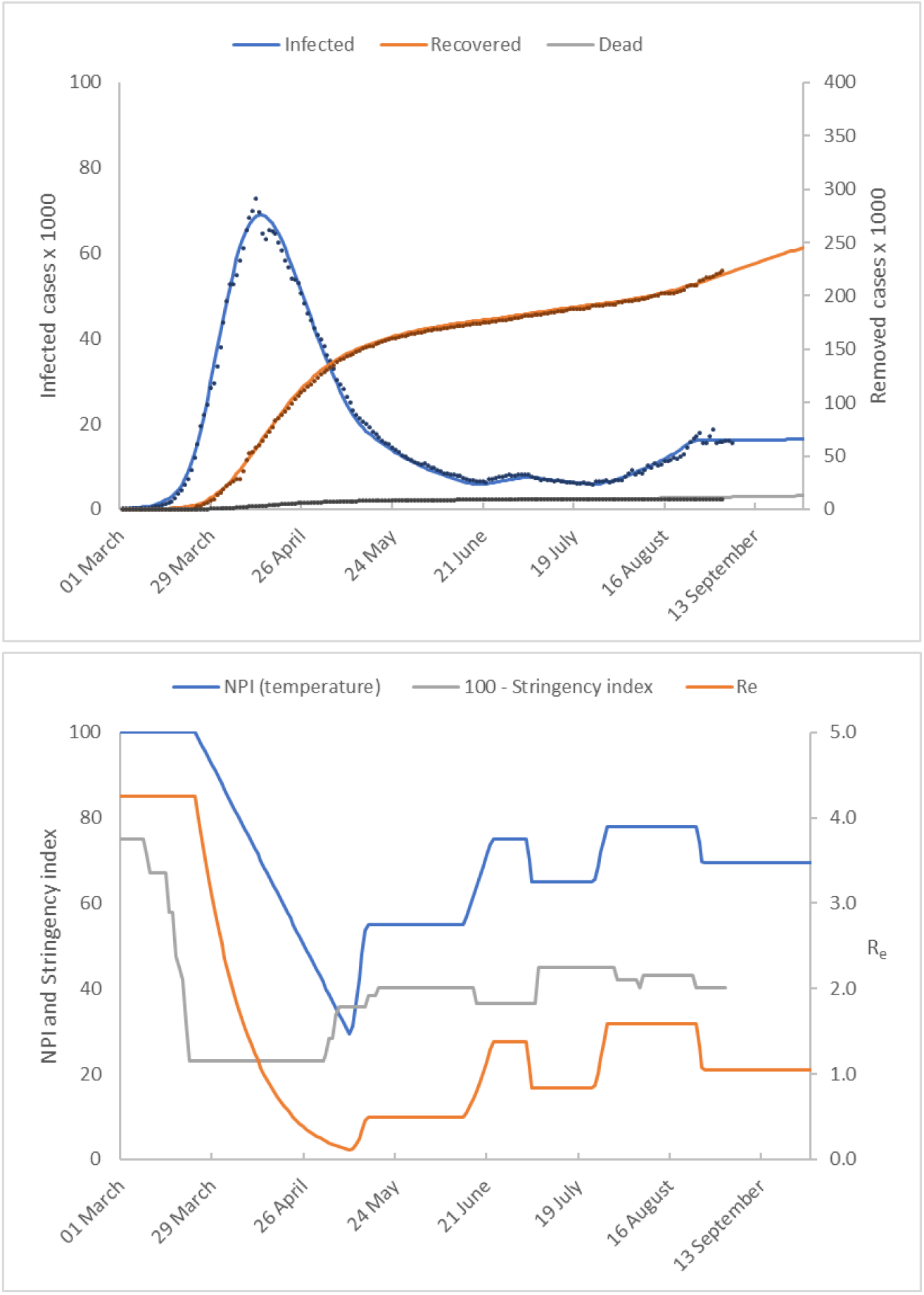
Germany. The upper chart shows the number of active infected, recovered and deceased cases as a function of time. Model predictions are shown as solid lines and reported values as discrete points. The lower chart shows the temperature profile representing NPIs imposed to achieve the model predictions. We compare this to the stringency index (plotted as 100 – Stringency Index). The estimate of the effective reproduction number is also shown.

The number of active cases peaked in mid-April and then declined. The introduction of Germany’s lock-down measures in late March is represented in our model by significant downward ramp in temperature over a 2-month period from the initial value of 100°C to around 30°C. This gives accurate model calibration of the early epidemic cases beyond the peak in infected cases up to 20 May when lock-down measures were relaxed.

The number of infected cases continues to decrease at a reduced rate until late June when a gradual increase commences. We capture this dynamic by extending the temperature profile progressively to maintain accurate model predictions. The first increase in temperature captures the reduced rate of decline in infected cases from relaxation of NPIs on 20 May. We find that the measures result in a sharp increase in *R_e_* followed by a sustained period where remains at this new level until mid-June 2020. The second temperature increase corresponds to the slight increase in infected cases reported from mid-June 2020. The final temperature increase on 23 July captures the increasing infection rate through to late August. Some NPIs were reintroduced on 31 July with further measures introduced on 23 August. This is captured in our model by a downward temperature ramp on 25 August.

The calibrated model constants obtained were as follows

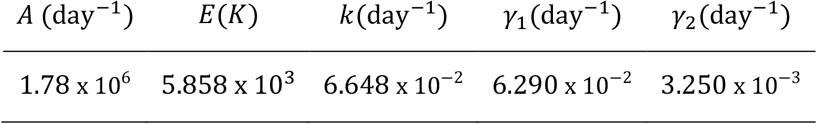

Using these calibrated model constants, we calculate R_0_ = 4.0 We observe an increase in the number of active cases in mid-June 2020, and that and that our model predicts *R_e_* > 1. In addition, there is a small downward trend in the number of active cases at the end of June 2020. This is captured in our model by a further decrease in temperature with a corresponding fall in *R_e_* to a value less than one. In late July there is a growing increase in the number of active cases which continues until 24 August during which time the model predicts an effective reproduction number of, *R_e_* = 1.5. By the end of August the model predicts a decrease in *R_e_* to 1.05.

Finally, assuming no additional NPIs and *R_e_* remains constant at a value of 1.05 the model is used to simulate the increase in infection rate for a further 28 days. Under these conditions the model forecasts virtually no change in the level of infected cases throughout September.

Interestingly, the final temperature profile shows a similar trend to Oxford’s government stringency index (shown in Figure 2 as ‘100 – stringency index’), [21]. The stringency index is an aggregate measure of governments’ responses to the COVID-19, which includes a variety of diverse measures for example school closures, travel bans etc. We suggest that the disparity between our temperature profile and the stringency index could arise for a number of reasons. It may be due to the relative weightings used to aggregate terms in the stringency index, the relative adherence of a population to introduced measures or the dynamics of the disease transmission rate.

### 3.2 Austria

Figure 3 shows *SIRD* model predictions (***R*_squared_** = 96.8%) plotted with the reported values for cumulative numbers of active infected, recovered and deceased individuals. The time shifts for reporting delays are *t_d_I__* = 0, *t_d_R__* = 9 *days*, *t_d_D__* = 9 *days*.

**Figure 3.**
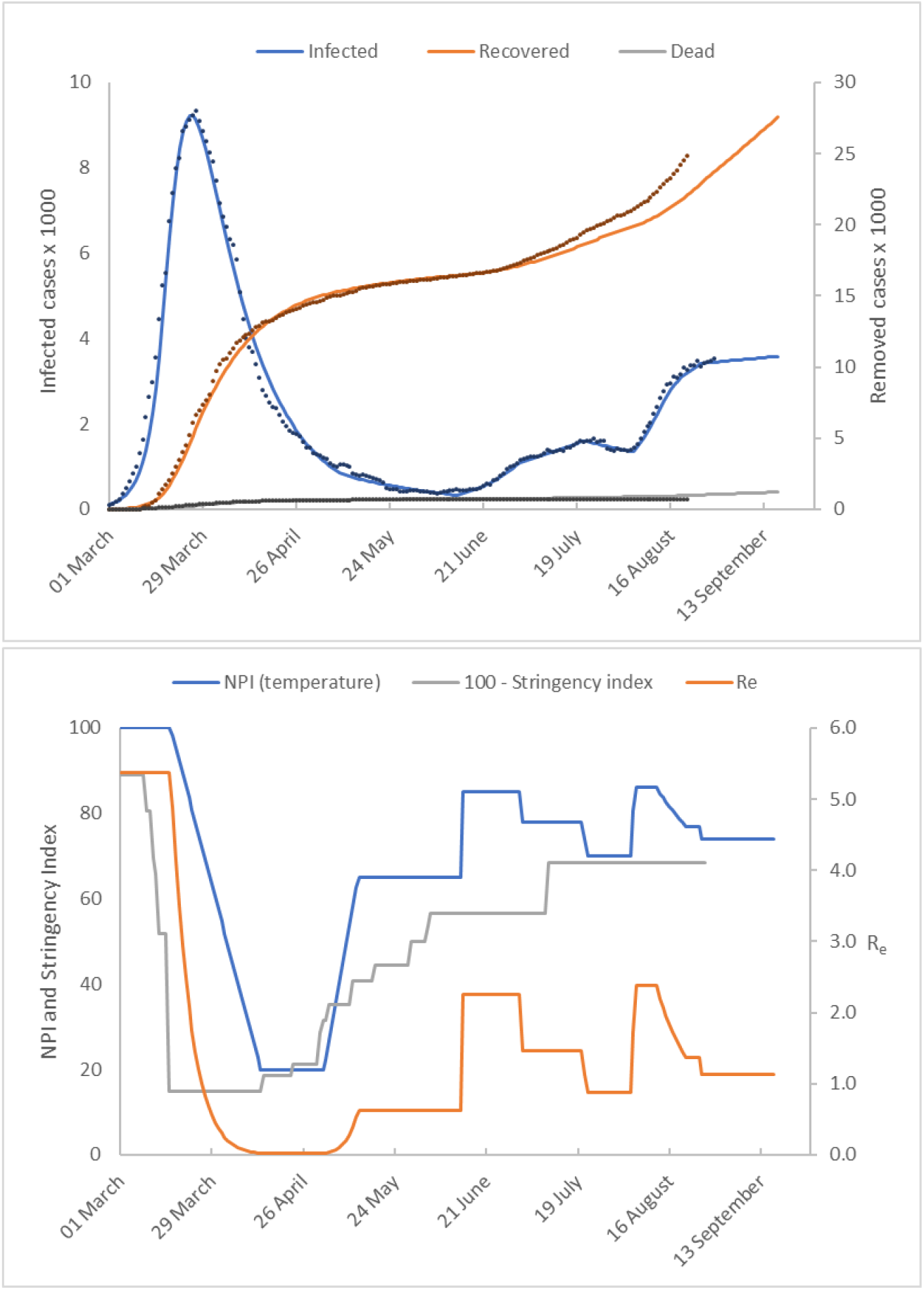
Austria. The upper figure shows the number of active infected, recovered and deceased cases as a function of time. Model predictions are shown as solid lines and reported values as discrete points. The lower figure shows the temperature profile representing NPIs imposed to achieve the model predictions. We compare this to the stringency index (plotted as 100 – Stringency Index). The estimate of the effective reproduction number is also shown.

The trajectories for the numbers of cases are similar to those shown in the German data. Austria successfully ‘flattened-the-curve’ of infected cases as early as the end of March, although the numbers of cases are an order of magnitude smaller than those reported by Germany. After successfully flattening the curve, NPIs were gradually relaxed and the number of active cases initially continued to decrease. In mid-June 2020, there was a gradual increase in the number of cases. In early August, a much steeper increase in the numbers of infected individuals occurs before the curve is flattened.

In our model, we first imposed a deeper and steeper downward temperature ramp taking the system temperature from 100°C to 20°C over a period of around 1 month.

The number of infected cases continues to decrease at a reduced rate until mid-June when numbers start to increase. We capture this dynamic by extending the temperature profile progressively to maintain accurate model predictions. The first increase in temperature captures the reduced rate of decline in infected cases from relaxation of NPIs in May. *R_e_* increases sharply, followed by a sustained period where *R_e_* remains at this new level until mid-June 2020. The second temperature increase corresponds to the slight increase in infected cases reported from mid-June 2020 after which the temperature is reduced to reflect the decrease in infected cases from 27^th^ June until 4^th^ August. The number of cases started to increase rapidly from this time. This is captured by a steep increase in temperature. The increase in cases is brought under control after that date. This is captured in our model by further decreases in temperature commencing on 13 August. Again, the temperature profile used in our model shows a similar trend to Oxford’s government stringency index.

The calibrated model constants obtained were as follows

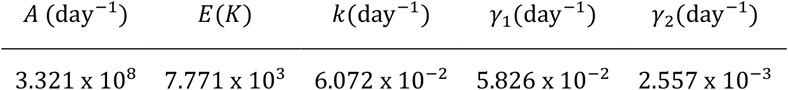

We calculate R_0_ = 5.37 followed by a fall in *R_e_* to a value *R_e_* < 1. We then observe an increase in the number of active cases starting in mid –June and that our model predicts an effective reproduction number of, *R_e_* = 2.38 by the 9^th^ August. This falls to *R_e_* = 1.13 by the end of August. If no additional NPIs were introduced and *R_e_* remained at this value, our model forecasts a small increase in infected cases by late September.

### 3.3 Saudi Arabia

Figure 4 shows *SIRD* model predictions (***R*_squared_** = 99.24%) plotted with the reported values for cumulative numbers of active infected, recovered and deceased individuals. The time shifts for reporting delays are *t_d_I__* = 0, *t_d_R__* = 10 *days*, *t_d_D__* = 10 *days*.

**Figure 4.**
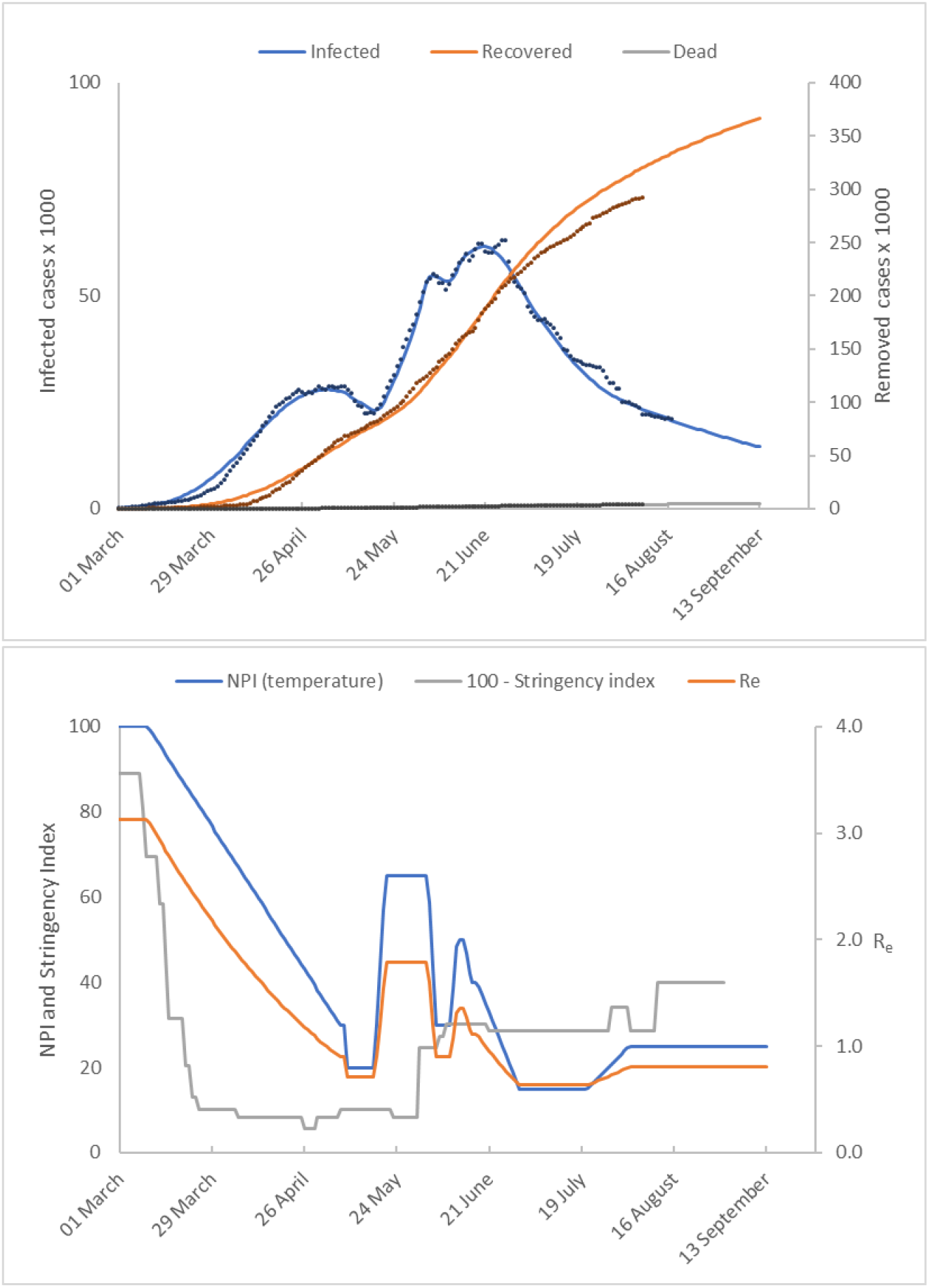
Saudi Arabia. The upper figure shows the number of active infected, recovered and deceased cases as a function of time. Model predictions are shown as solid lines and reported values as discrete points. The lower figure shows the temperature profile representing NPIs imposed to achieve the model predictions. We compare this to the stringency index (plotted as 100 – Stringency Index). The estimate of the effective reproduction number is also shown.

Initially, the trend in cases is similar to Germany and Austria. The number of infected cases is successfully curtailed by early May. The initial temperature ramp reduces the system temperature from 100°C to 30°C over a period of around 6 weeks. The model predicts a much slower response *R_e_* in to the initial NPI when compared to Germany and Austria. This is reflected in the calibrated values of the coefficients *A* and *E* in equation (10).

To fit the reported data for the rapidly increasing numbers infected cases it was necessary to impose a sharp temperature increase on 18 May which raises *R_e_* to 1.5. This was followed by a series of step changes in temperature which were introduced to replicate the pattern
 manifest in the reported data. We calculate R_0_ = 3.3 and an effective reproduction number of, *R_e_* = 0.8 throughout August. Assuming these conditions are maintained the model forecasts a continuing decline in infected cases throughout September.

The calibrated model constants that we obtained were as follows,

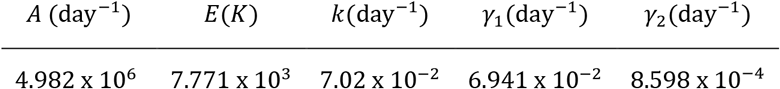

### 3.4 Italy

Figure 5 shows *SIRD* model predictions (***R*_squared_** = 96.7%) plotted with the reported values for cumulative numbers of active infected, recovered and deceased individuals. The time shifts for reporting delays are *t_d_I__* = 0, *t_d_R__* = 10 *days*, *t_d_D__* = 10 *days*.

**Figure 5.**
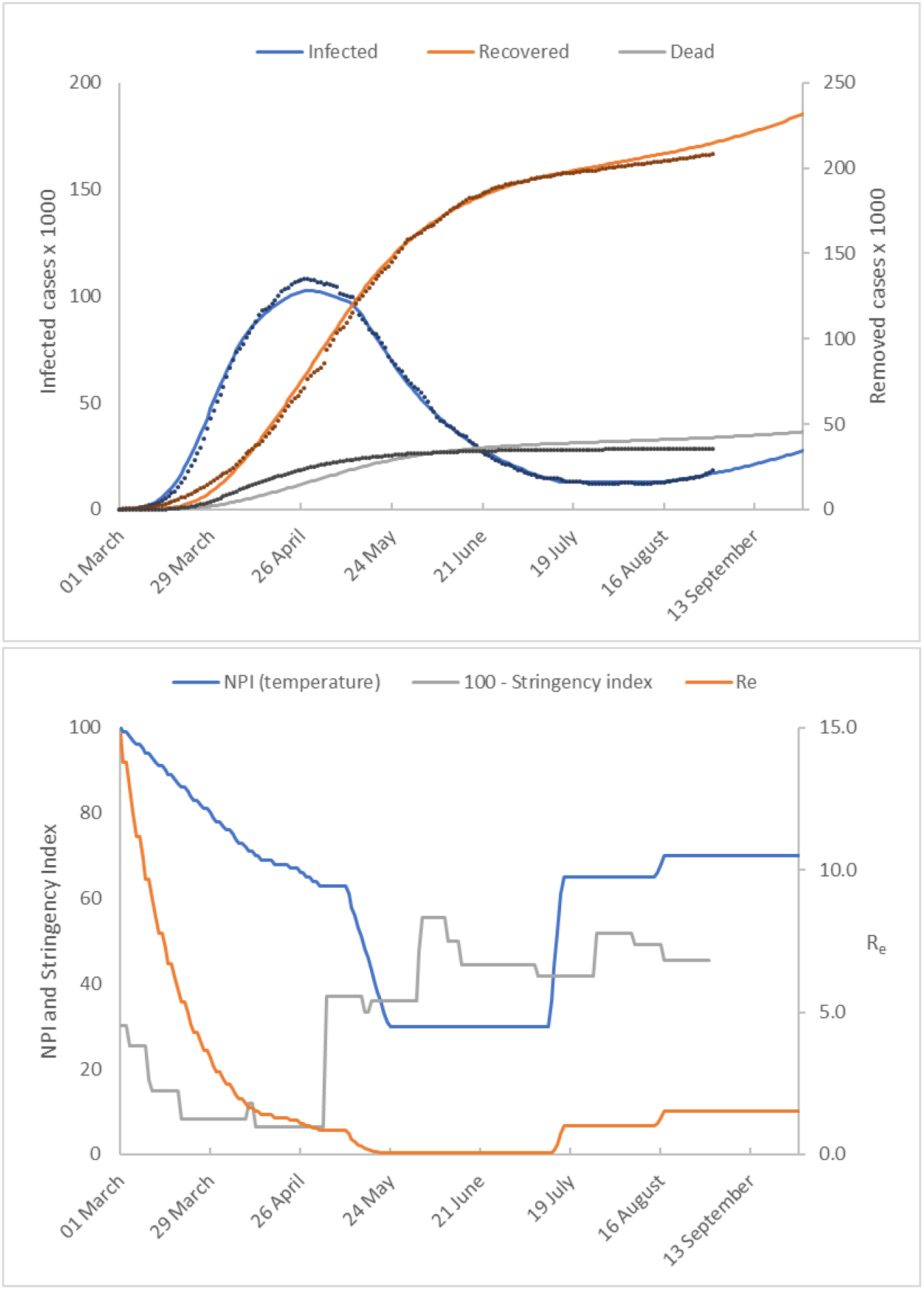
Italy. The upper figure shows the number of active infected, recovered and deceased cases as a function of time. Model predictions are shown as solid lines and reported values as discrete points. The lower figure shows the temperature profile representing NPIs imposed to achieve the model predictions. We compare this to the stringency index (plotted as 100 – Stringency Index). The estimate of the effective reproduction number is also shown.

The trend in cases is similar to the pattern in data from Germany and Austria. Italy successfully ‘flattened-the-curve’ of infected cases by the end of April, and infections and deaths declined steadily. A temperature decrease is required after the introduction of the most stringent of Italy’s lock-down measures in late February. This takes the form of a downward ramp, lasting over a 2-month period corresponding to a fall in the initial temperature of 100°C to a temperature of around 60°C.

Italy first began to relax NPIs around the beginning of May 2020, however in order to calibrate the model a further reduction in temperature was required to a constant value of 30°C. In mid July the reported number of infected cases becomes constant and remains so until mid August when it starts to increase. We capture these dynamics first by a sharp temperature increase which raises *R_e_* to 1.0 and then by a second increase in temperature which raises *R_e_* to 1.5. Using these conditions the model forecasts that the number of infected cases will increase to 27000 by late September.

The calibrated model for Italy is reasonably good, however there is a significant model data mismatch for the number of the dead. What is strikingly different about the model for Italy is the prediction that R_0_ = 15 and that the value of *k* is only about half that of the other countries. We can offer no explanation for these results other than to say that the modelling is based on a rigorous mass balance of all model elements and is therefore affected by errors in data. We note that on 4 August the international news media reported that Italy announced that the actual number of infections was estimated to be six times higher than the reported number which may be a factor in these questionable model predictions.

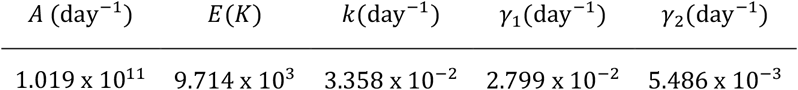

## 4.0 Discussion and conclusions

The primary aim of this work is to demonstrate the effectiveness of using parameter regression methods to calibrate an *SIRD* model for COVID-19 where the effective reproduction number response to NPIs is non-linear and variable in terms of response rates, magnitude and direction. By using an existing commercial chemical engineering package capable of parameter regression with piecewise continuous integration with event and discontinuity management we have been able to explore the efficacy of this approach. We have highlighted the trend in the number of active cases in Germany, Austria and Saudi Arabia and Italy.

Results indicate that our model where *R_e_* varies exponentially as a function of NPIs can accurately capture the reported numbers of disease progression in the sample of countries selected. We elected to keep our mathematical model comparatively simple using the established *SIRD* scheme. Our enhancements to this scheme are that we treat *R_e_* as a variable stoichiometric coefficient, and assume that it varies exponentially. We have used temperature as a placeholder to develop this exponential variation. This choice is expedient as the modelling platform already had an Arrhenius equation model with regression tools. A benefit of this approach is that a non-linear response in *R_e_* to NPIs is transformed to a linear ramp. This transformation made it easier to calibrate the model using a systematic series of manual interventions. The experience gained in using this approach for several data sets suggests that a strategy for model calibration may be developed into an algorithm which could be coded.

Understanding that in an *SIRD* model *R_e_* is a variable stochiometric coefficient in the infection step has enabled the determination of *R_e_* ▪ *k* by model calibration together with their numerical decoupling. This has resulted in identifying the characteristic time for *k* to be in the range of 14 to 16 days with Italy being an outlier at around 30 days.

The modelling software used in this study is designed for simulation studies of batch chemical systems. The software has limitations, not the least being the considerable expertise needed to utilise it for this application which is significantly outside its designed purpose, and we are not advocating its use. Rather we are suggesting that the methodology embedded in the software with some further development of regressive capability could be developed into an effective software tool for epidemic study. This work is merely a demonstrator of the algorithmic steps involved.

One aspect of the modelling software’s capability which was not used in this study is simulation of thermal runaway – the behaviour of a reacting system which accelerates exponentially. Batch chemical process development places strong emphasis on avoiding a violent thermal runaway, which a significant number of processes could potentially undergo unless properly designed. The methods for studying and the design procedures to negate the possibility of a thermal runaway are well established. There are many analogies in a thermal runaway scenario with the outbreak of an epidemic. For example, the explicit inclusion of the effective reproduction number in the model equation for infection reveals the stability characteristics of the system. Another feature which could be exploited is the modelling of reagent additions which is directly analogous to an influx of infected cases to a population. Such scenarios will be simulated and reported in a future publication.

In conclusion, this study has attempted to assess the potential of some established chemical engineering modelling principles and practice for application to modelling of epidemiological systems. We have successfully developed a novel extension to the analogy between chemical and epidemiological system models.

## Data Availability

Data used to develop models was obtained from public domain sources.

https://www.worldometers.info/coronavirus/

Competing interests

The authors declare that they have no competing interests.

## Footnotes

1 The simulator used for this study is BatchCAD 7.1. The software was originally developed by Bramfitt VJ, Wright AR and Wright AW from 1986 to 1999, and was eventually acquired by Aspen Technology Inc.

2 Model equations for chemical systems are more commonly expressed in terms of concentration. Epidemiological models are expressed in terms of population fraction. The basis of the model equations for chemical schemes can be transformed from concentration to mole fraction by considering molecular weight and density. It can be shown that by assuming all molecular weights and densities have equal value, the numerical values of mole fraction and concentration are identical, and the model equations are equivalent.

3 The simplex algorithm is modified and uses intelligent jacketing which exploits embedded knowledge of the characteristics of kinetic constants in chemical reaction systems to improve robustness and aid convergence.

